# Application of transformers for predicting epilepsy treatment response

**DOI:** 10.1101/2020.11.10.20229385

**Authors:** Jiun Choong, Haris Hakeem, Zhibin Chen, Martin Brodie, Nicholas Lawn, Tom Drummond, Patrick Kwan, Zongyuan Ge

**Affiliations:** Monash University, Melbourne, Australia; Alfred Hospital, Melbourne, Australia; University of Glasgow, Glasgow, Scotland; WA Adult Epilepsy Service, Perth, Australia

**Keywords:** D eep learning, transformer, self-attention mechanism, epilepsy, treatment, machine learning, prediction, time-series, longitudinal, prediction

## Abstract

There is growing interest in machine learning based approaches to assist clinicians in treatment selection. In the treatment of epilepsy, a common neurological disorder that affects 70 million people worldwide, previous research has employed scoring methods generated from traditional machine learning methods based on pre-treatment patient characteristics to classify those with drug-resistant epilepsy (DRE). In this study, we used an attention-based approach in predicting the response to different antiseizure medications (ASMs) in individuals with newly diagnosed epilepsy. By applying a conventional transformer to model the patient’s response, we can use the predicted probability to determine the success rate of specific ASMs. Applying the transformer allowed the model to place attention on patient information and past treatments to model future drug responses. We trained a conventional transformer model based on one cohort of 1536 patients with newly diagnosed epilepsy, compared its performance with other trained models using RNN and LSTM, and applied it to a validation cohort of 736 patients. In the development cohort, the transformer model showed the highest accuracy (81%) and AUC (0.85), and maintained similar accuracy and AUC (74% and 0.79, respectively) in the validation cohort.

## 1. INTRODUCTION

Epilepsy is one of the common neurological diseases affecting approximately 0.8% of people during their lifetime.^1^ It is characterized by an increased tendency to have recurrent seizures at unpredictable times which can lead to physical injury, impair social functioning, affect mental health and quality of life or even cause death.^2^ Despite advances in non-pharmacological treatment options over the years in the form of resective surgery, neuromodulation and dietary therapies, the mainstay and first-line treatment remains as drug therapy, for which over 20 antiseizure medications (ASMs) are currently available. However, drug selection still relies on a trial-and-error approach and 30% of patients have drug-resistant epilepsy (DRE) that does not respond to currently available ASMs.^3^ Although there are general guidelines based on broad seizure types, there is currently no reliable way to predict the optimal drug choice for individual patients.

There is growing interest in applying machine learning in assisting healthcare decision making, fuelled by advancements in the deep learning field such as BioBERT for natural language processing (NLP) tasks,^4^ generative adversarial networks (GANs) for medical image generation^5^ and convolutional neural networks (CNNs) for detection through image classification.^6^ In epilepsy, the application of machine learning has been limited to traditional models such as random forest (RF) algorithms^7^ and scoring based systems^8^ to predict a patient’s ASM treatment outcome. Furthermore, recent advances with transformers which uses attention-mechanisms have proven success in many domains^4, 9, 10^ due to the ability to focus on specific parts of the data.

In this study, we applied an attention-based model to predict the optimal ASM prescription for individual patients based on a broad range of demographic factors and epilepsy information. The performance and robustness were verified through a separate cohort that was not used in the training or validation of the transformer model.

Our contributions are as follows:

- We are the first to utilise and show that an attention-based model can capture the latent structure for individual epilepsy patient response to ASM treatment. The transformer model is able to capture additional information from the demographic and ASM information due to the multi-headed self-attention mechanism.
- We show that the transformer model is more suitable for capturing the relevant longitudinal information in predicting treatment response compared to other time-series based models.
- We achieve reasonable performance with an external validation cohort in Section 4.4 without further fine-tuning of the model. The transformer model is able to generalise to an extent as seen in Section 4.5.

## 2. RELATED WORK

Machine learning in epilepsy management can potentially assist epileptologists in clinical decision making in diverse domains such as automated analysis of electroencephalography (EEG) and diagnosis though images, prediction of responses to ASM, and resective surgery.^11^ Past papers that investigated epilepsy treatment typically used traditional ML algorithms^7^ and there has been a recent shift towards deep learning ML algorithms due to their exceptional performances in different domains.^12^ Other cases of modelling within epilepsy is to predict seizure recurrence after withdrawal from ASMs by using a scoring system.^8, 13^ These models generally include demographic and clinical risk factors identified by standard statistical methods. There has also been a recent paper that employed a manually tuned mathematical model to determine which ASM to use.^8^ A few studies have also looked at developing individualized prediction models for early diagnosis of DRE^14, 15^ and for selecting the most appropriate ASM.^6, 14, 15^

In recent advances of deep learning, attention-based models have shown promise in various applications such as NLP, image recognition, and even electronic health records (EHR) data.^4, 10, 16^ Attention-based models is a method in deep learning that allows the network to break down complex inputs into smaller parts for a sequence of data. The backbone of transformers utilises these attention-mechanisms in a modular fashion to focus its attention on different parts of the data.^17^ Although there have been applications of transformers within the medical domain, the usage of transformers to predict treatment outcomes has not been adapted before.

## 3. METHODS

In this section, we will discuss the methodology applied to train and validate the model. Due to the nature of longitudinal data encompassing a time aspect, time-series models are used for baselines to compare against the transformer model. Recurrent neural networks (RNN) and long-short term memory cells (LSTM) are networks that are used to compare against the transformer. Figure 1 depicts the high level overview of the procedure used for using the transformer to predict ASM treatment response for patients.

**Figure 1.**
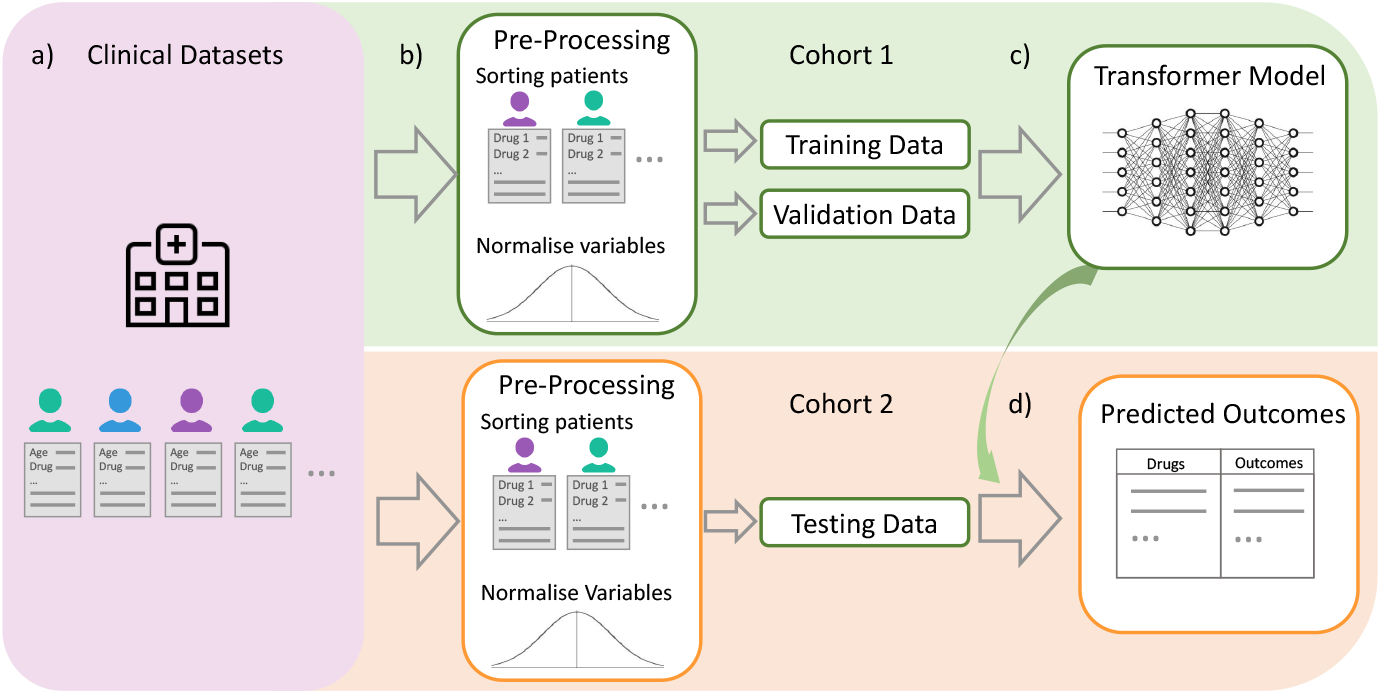
High level overview of using the transformer model for longitudinal data. a) Each row of patient data corresponds to an ASM taken for a duration of time. Individual patients may trial different ASMs. b) The raw data is normalized across each variable to *µ* = 0 and *σ* = 1 within the training set and applied to the validation and test set. ASM trials are grouped by patient and split into training/validation sets. c) The transformer is trained and tuned with the first cohort d) The transformer will output a treatment outcome probability for each patient based on the ASM provided. The model can provide a prediction to the ASM treatment response for individual epilepsy patients.

### 3.1 Baseline models and transformer

This section explains the structure of the baseline models and the transformer architecture. Figure 2 highlights the different structures of each model.

**Figure 2.**
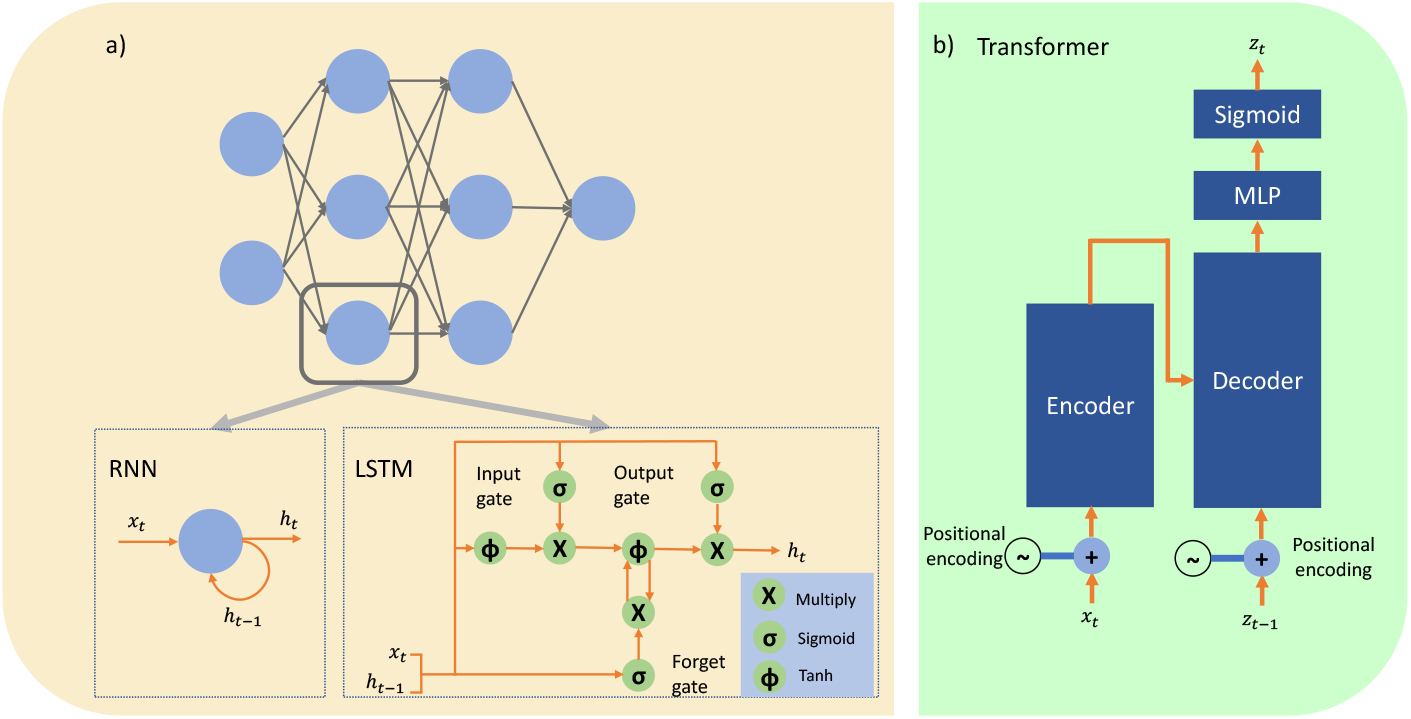
In both figures, *x*_*t*_, *h*_*t*_, *h*_*t−*1_, *zt* represents input, output from current neuron, output from the previous time period of the current neuron, and output at the final layer respectively. a) The top shows the common MLP connection for a RNN and LSTM network. RNN cells have a direct recurrent connection to itself. LSTM cells have several gates to control the long and short term memory of the inputs and previous outputs. b) High level overview of the transformer

#### 3.1.1 RNN

Recurrent neural networks (RNN) have a feedback aspect within the neuron to roll back input sequences for time-series data.^18^ Basic RNNs follow a standard multi-layer perceptron (MLP) except for each neuron in the hidden layer, there is a connection that feed back into itself. Hence, this weight can be updated and it allows the neuron to retain temporal information for a sequence of data. However, due to the nature of a RNN having a simple recurrent loop, the network cannot hold long term memory as the gradient flowing back in time will either diminish or explode.^18^

Before the prevalence of transformers, RNNs were typically used for time series data due to the simplicity of the network. However, RNNs are difficult to train and extending the complexity of the network results in vanishing and exploding gradients during back propagation of the network.^19^

#### 3.1.2 LSTM

LSTMs tackles the long term dependency by introducing “gates” into the cell where the gates act as a switch to control the reading and writing of inputs within the cell.^18^ As the gates control the read and write of past input sequences, it mitigates the vanishing and exploding gradients problem encountered by a RNN. LSTMs are more complex compared to a basic RNN, as it has several gates and additional activation functions within the model.^18^

Although LSTMs were created to mitigate the vanishing and exploding gradient problems faced by RNNs, LSTMs are unable to model extensively long term dependencies. Furthermore, LSTMs are difficult to train in nature compared to a non-recurrent neural network.

### 3.2 Transformer model

The transformer model is a relatively new deep learning model that uses an attention mechanism which allows the model to pay attention to different parts of an input sequence of data.^17^ Transformers are able to focus on different parts of the data by dividing the self-attention mechanism into parts and aggregating the results in the latter part of the model.

In this work, we will apply a vanilla transformer^17^ to predict ASM treatment responses at the individual patient level, using longitudinal registries of new onset epilepsy patients containing comprehensive diagnostic and follow-up information. Transformers have demonstrated prominence in modelling of longitudinal electronic health records (EHR) for personalised diagnosis.^16, 20^ We adopt the transformer to predict individual patient response to ASM treatment with longitudinal data.

To adapt the transformer to the longitudinal data, each ASM trial is a separate input to the transformer as seen in Figure 3. The inputs are passed on sequentially for each patient with the inputs being reset when it reaches the next patient. The inputs are first passed through a positional encoder to encode the time step for the ASM trial. The encoded data is passed through a multi-head attention layer described as:

**Figure 3.**
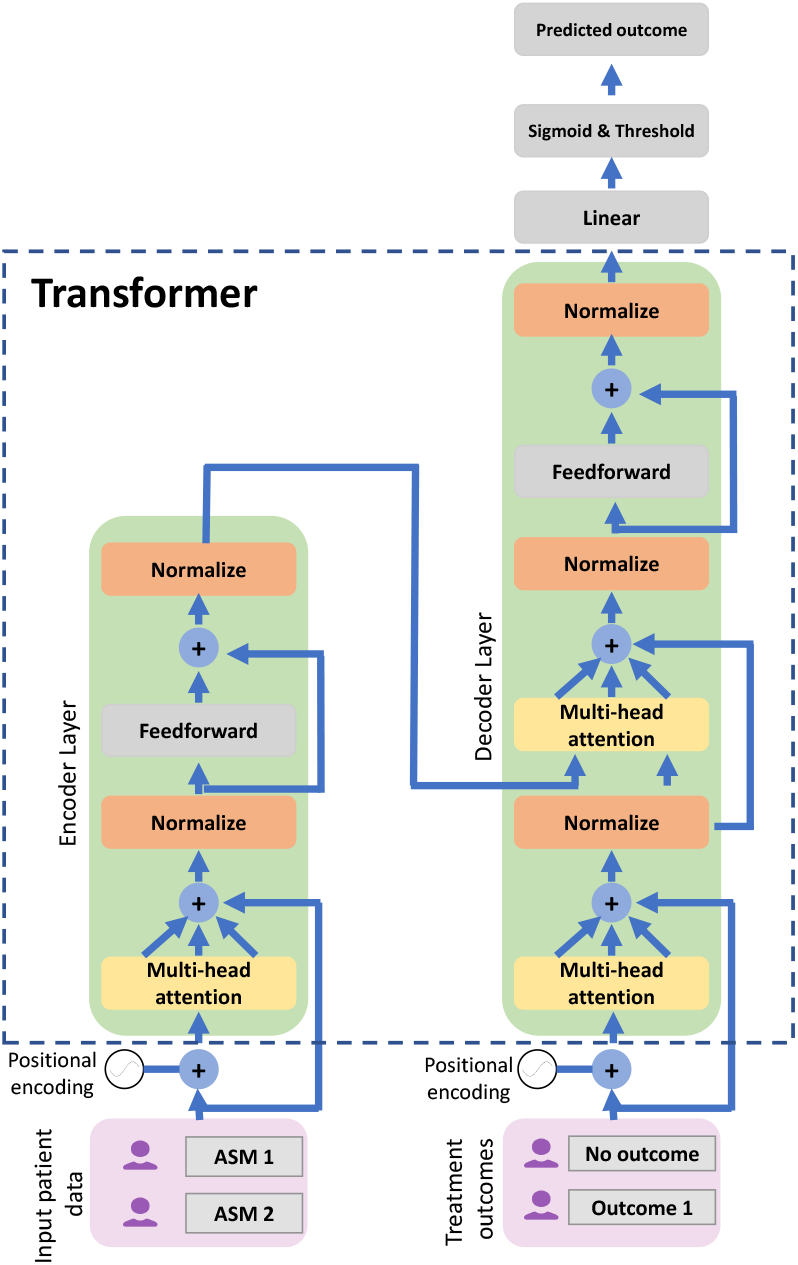
Overview of a single encoder-decoder pair for a transformer network. Note that the modularity of the transformer arises from stacking encoder and decoders. The transformer model utilises the original structure^17^ except there is no input embedding that needs to be learnt. Each ASM trial is fed into the network sequentially and the time period is encoded via the positional encoder.

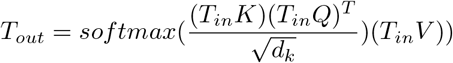

where *T*_*in*_, *T*_*out*_ are input and output data in the mult-head attention layer. *K, Q, V* are key, query, and value vectors that retrieves corresponding values for each of the inputs. 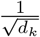 represents a scaling factor for the queried value. The multi-attention layer analyses the encoding of the current ASM trial and its encoded information such as demographic and past ASM trials, and relates it to other ASM trials. There are several multi-head attention layers within the network as the transformer builds its own representation of the data within the feedforward layers. The inputs to the decoder are the treatment responses from previous ASM trial outcomes and current ASM trial information passed from the encoder.

We utilise the transformer in this setting due to its ability to capture hidden latent dependencies between each patient ASM trial, and the ability of the attention mechanism to focus on different ASM treatment outcomes. The transformer model also allows a variable length of patient visits, hence it is not restricted to only training and validating the model on the first few regimen. This allows the transformer model to effectively utilise the entire dataset.

### 3.3 Model training

To utilise the data effectively, the model was trained on each progressive ASM trial. For example, a patient treated with 2 regimens will be used to train the model 2 times, with the first time only using the first regimen and the second time utilising both ASM trials. Furthermore, the final treatment outcome for the current sequence of data is always masked within the training data to avoid the model from seeing the current treatment outcome. The transformer model uses a 5-fold cross validation approach on the Glasgow dataset to obtain optimal hyper-parameters. Table 4 in the Appendix shows the final hyper parameter values that were used. The model was used to validate its performance on the Perth cohort without any further training of the models using the Perth dataset as seen in section 4.5.

## 4. EXPERIMENTS

### 4.1 Dataset

The data was obtained from two longitudinal registries of patients maintained in UK (Glasgow) and in Australia (Perth). These registries include a total of 2,272 adults with newly diagnosed and treated epilepsy. The Glasgow dataset (n=1536) includes patients seen at the Epilepsy Unit of Western infirmary in Glasgow, Scotland from 1 July 1982 to 31 October 2012 and followed up prospectively till 30 April 2016 or death. The Perth dataset (n=736) has started registering patients seen at First Seizure Clinics in Western Australia since 1999 and is being actively maintained. The patients’ characteristics, treatment approach, and database structure are similar across the registries. The registries collect similar patient data including demographics, medical history, family history, epilepsy risk factors, ASM regimens, pre-treatment seizure number and frequency, EEG and MRI results, and treatment response (including seizure control and tolerability).

Between the two cohorts, there are differences in the distribution with the demographic information such as the patient history, EEG, age initiated and MRI results as seen in Figure 4. The Perth dataset is more dispersed across the age group whereas the Glasgow dataset has a younger population. There is also a lower proportion of epileptogenic abnormalities on brain imaging and higher proportion of patients with unknown aetiology in the Glasgow cohort which could be due low field MRI scans before the year 2000. The list of variables describing the patients used in this work* can be found in table 5.

**Figure 4.**
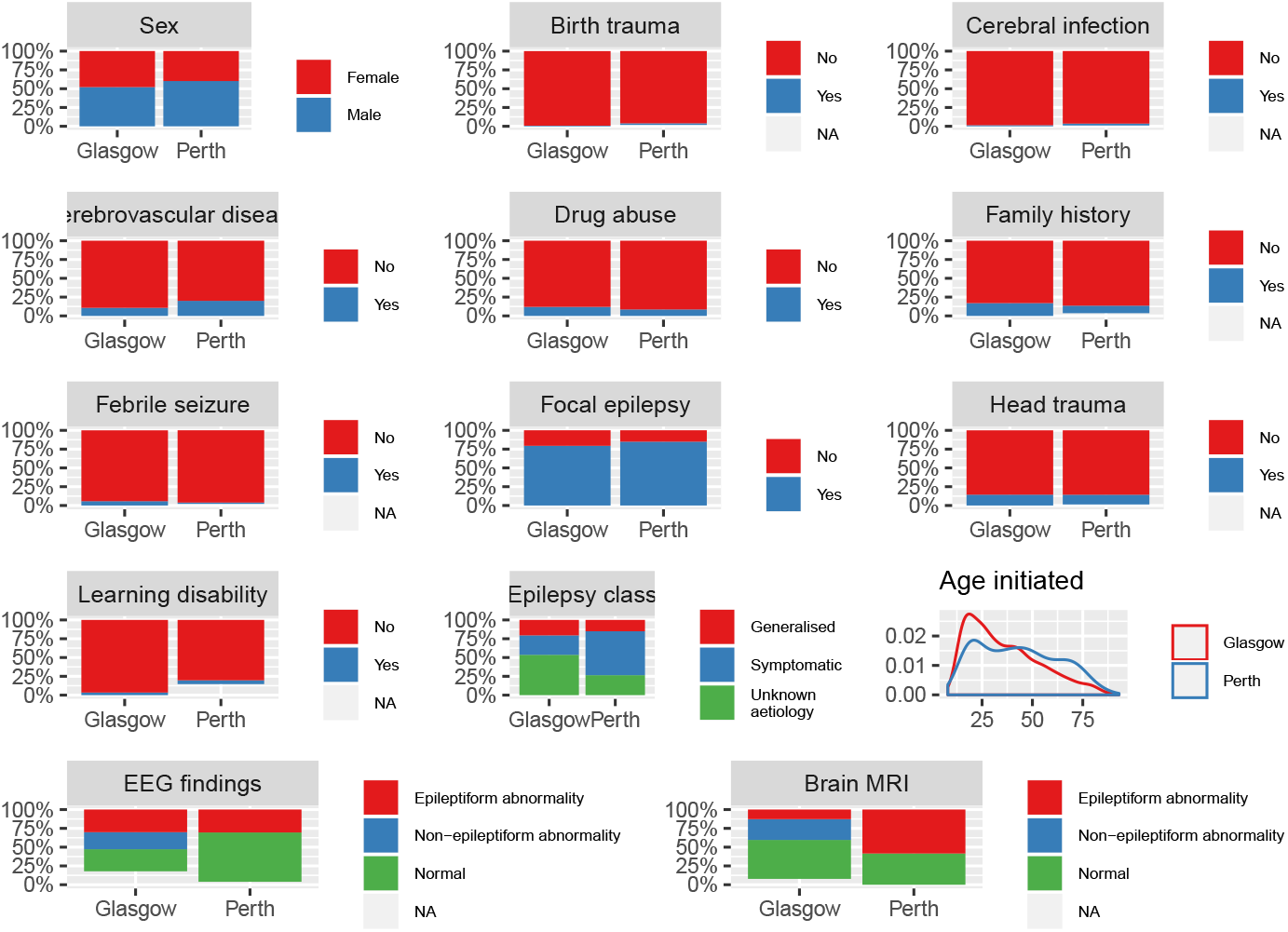
Side-by-side comparison of the Glasgow and Perth cohort patient data. The y-axis represents the % of patients within that cohort with the coloured label.

### 4.2 Pre-processing

The dataset was normalized before being used for training and validation of the model. Before normalization, the categorical variables were binarised into separate columns for each category and continuous variables were split into discrete categories to reduce the complexity for the model. Furthermore, ASMs that were not commonly prescribed were removed from both cohorts.

To prevent information from leaking between the training and validation cohorts, the normalization step was calculated from the training samples for each input variable (*µ*_*i*_ = 0, *σ*_*i*_ = 1, *where i is the i*^*th*^ *input variable*) and the normalization values were applied to the validation samples. This allows the transformer model to predict treatment outcomes of new patients by applying the same normalization. Moreover, the complexity of the input variables were further reduced by categorising continuous variables into discrete categories based on quartiles. This was shown to improve the model performance as shown in Section 4.3.

### 4.3 Ablation studies

In this section, we perform ablation studies on the techniques described in Section 4.2 to isolate their individual contributions. The studies were performed with a 5-fold cross validation with the Glasgow cohort. Accuracy is determined by the predicted treatment outcome by the model based on each ASM trial, with a threshold of 0.5 to separate successful and unsuccessful treatment outcomes. This predicted treatment outcome is compared with the actual result from the ASM trial, and the accuracy is calculated across each ASM trial in the 5-fold cross validation. AUC is derived from the predicted treatment outcome values compared to the ground truth (0 = unsuccessful, 1 = successful) as the model predicts values between 0 to 1.

#### Categorising continuous variables

To reduce the complexity of the data, continuous variables such as age were categorised into equal segments. The results in Table 1 highlights the effectiveness of reducing the complexity of the input data by discretizing the continuous variables.

**Table 1.**
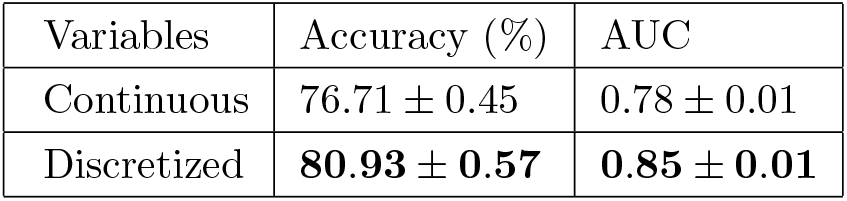
Discretizing continuous variables. Numbers represent (*µ ± σ*)

#### Multi-head attention

Here, we study the effects of tuning the number of heads for the multi-head attention mechanism. The results in Table 2 highlight the use of additional heads in improving the performance of the model. This shows that the model benefits from separating the embedding spaces within the network.

**Table 2.** Effect of multi-head attention. Numbers represent (*µ ± σ*)

| # of heads | Accuracy (%) | AUC |
| --- | --- | --- |
| 1 | $80.54 \pm 0.36$ | $0.84 \pm 0.01$ |
| 2 | **$80.93 \pm 0.57$** | **$0.85 \pm 0.01$** |
| 3 | $80.53 \pm 0.60$ | $0.83 \pm 0.01$ |

### 4.4 Results

The model performances were based off a 5-fold cross validation split of the Glasgow dataset. Table 3 highlights the capability of the transformer to model the variable lengths between epilepsy treatment outcomes. In contrast, RNN and LSTM have trouble modelling the different time lengths across the patient dataset. LSTM converges to classifying a single treatment outcome for each ASM treatment while the RNN converges to a performance that is lower than the transformer model. This can also be explained with the sequences of data typically being around 2-4 trials long, with the highest number of trials within the dataset being 13 trials for an individual patient. Due to the nature of the dataset, the longer term dependencies are not as prevalent compared to the shorter term dependencies for each treatment response.

**Table 3.** Model testing performance comparison on Glasgow cohort. Numbers represent (*µ ± σ*)

| Model | Accuracy (%) | AUC |
| --- | --- | --- |
| RNN | $75.18 \pm 0.85$ | $0.78 \pm 0.02$ |
| LSTM | $75.83 \pm 0.73$ | $0.69 \pm 0.03$ |
| Transformer | **$80.93 \pm 0.57$** | **$0.85 \pm 0.01$** |

**Table 4.** Tuned hyper-parameters for the final transformer model

| Hyper-parameter | Final value |
| --- | --- |
| Number of encoder/decoder pairs | 3 |
| Number of heads | 2 |
| Learning rate | 3e-4 |
| Learning rate decay factor | 7e-1 |
| Early stopping epochs | 10 |
| Number of epochs | 50 |

**Table 5.** List of variables describing the patient.

| Variable | Description |
| --- | --- |
| Sex | Biological gender of the patient |
| Age initiated | Age at initiation with the first antiseizure medication treatment |
| Epilepsy class | Epilepsy classification |
| Focal | Dichotomised epilepsy classification, whether it is focal epilepsy or not |
| Family history | Prior to the treatment initiation, whether the patient has family history of epilepsy or not |
| Febrile | Prior to the treatment initiation, whether the patient has history of febrile seizure or not |
| Cerebral infection | Prior to the treatment initiation, whether the patient has history of cerebral infection or not |
| Birth trauma | Prior to the treatment initiation, whether the patient has history of birth trauma or not |
| Head injury | Prior to the treatment initiation, whether the patient has history of head injury or not |
| Drug | Prior to the treatment initiation, whether the patient has history of drug abuse or not |
| Alcohol | Prior to the treatment initiation, whether the patient has history of alcohol abuse or not |
| Cerebrovascular disease | Prior to the treatment initiation, whether the patient has history of cerebrovascular disease or not |
| Psychiatric comorbidities | Prior to the treatment initiation, whether the patient has history of psychiatric comorbidities or not |
| Learning disability | Prior to the treatment initiation, whether the patient has history of learning disability or not |
| Brain MRI | Categorized CT/MRI findings |
| EEG category | Categorized EEG findings |
| Outcome | Terminal treatment outcome, at the end of study period |

Overall, the performance of the transformer can be attributed to the architecture of the model, as it focuses on relevant embeddings within the network due to the multi-head attention mechanism. It is also able to capture the time aspect effectively without the direct use of recurrent connections compared to RNNs and LSTMs.

### 4.5 Cross-Cohort validation

To ensure that the model is generalising well, a cross-cohort validation was used to test the transformer model. The trained transformer model from the Glasgow dataset was applied directly to the Perth dataset without further fine-tuning of the model. The accuracy and AUC on the Perth dataset are 74.05% and 0.79 respectively. Figure 5 shows the similarities of the two cohorts captured by the encoder of the transformer but there are still distinct clusters of each cohort that the transformer cannot capture. Figure 6 shows the misclassifications (indicated by the blue points) are clustered closely with the correctly classified treatment outcomes. The cluster of Perth data towards the bottom right of the Figure 5 indicates that the model has not been able to generalise information from the Glasgow patients as there are clusters of Perth patients resulting in misclassifications of the Perth cohort towards bottom right of Figure 6. This indicates that the transformer model requires additional relevant variables and data to further separate the treatment outcomes within the latent space of the model.

**Figure 5.**
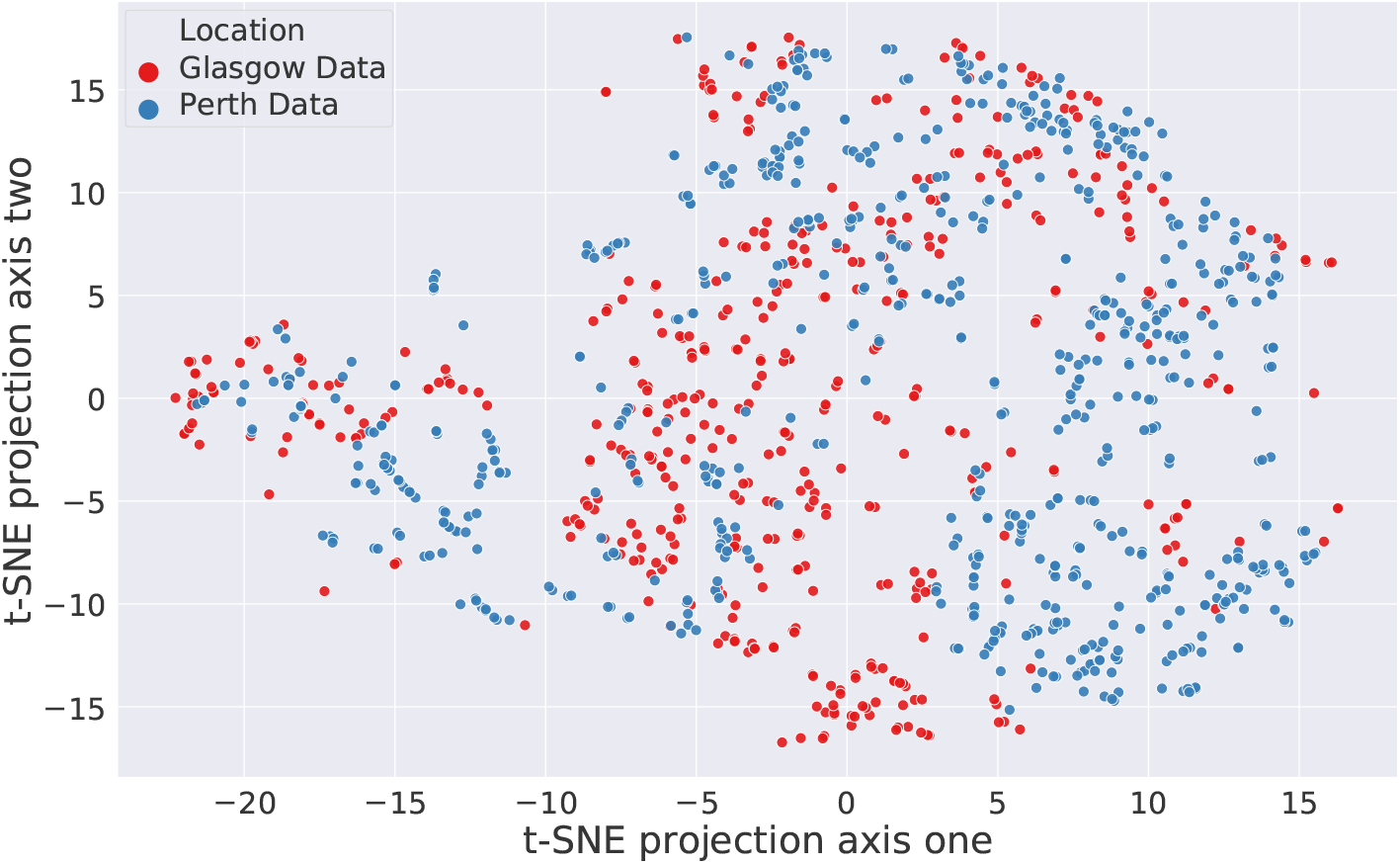
Projection of the Glasgow validation and Perth patients within the last encoder layer of the transformer network

**Figure 6.**
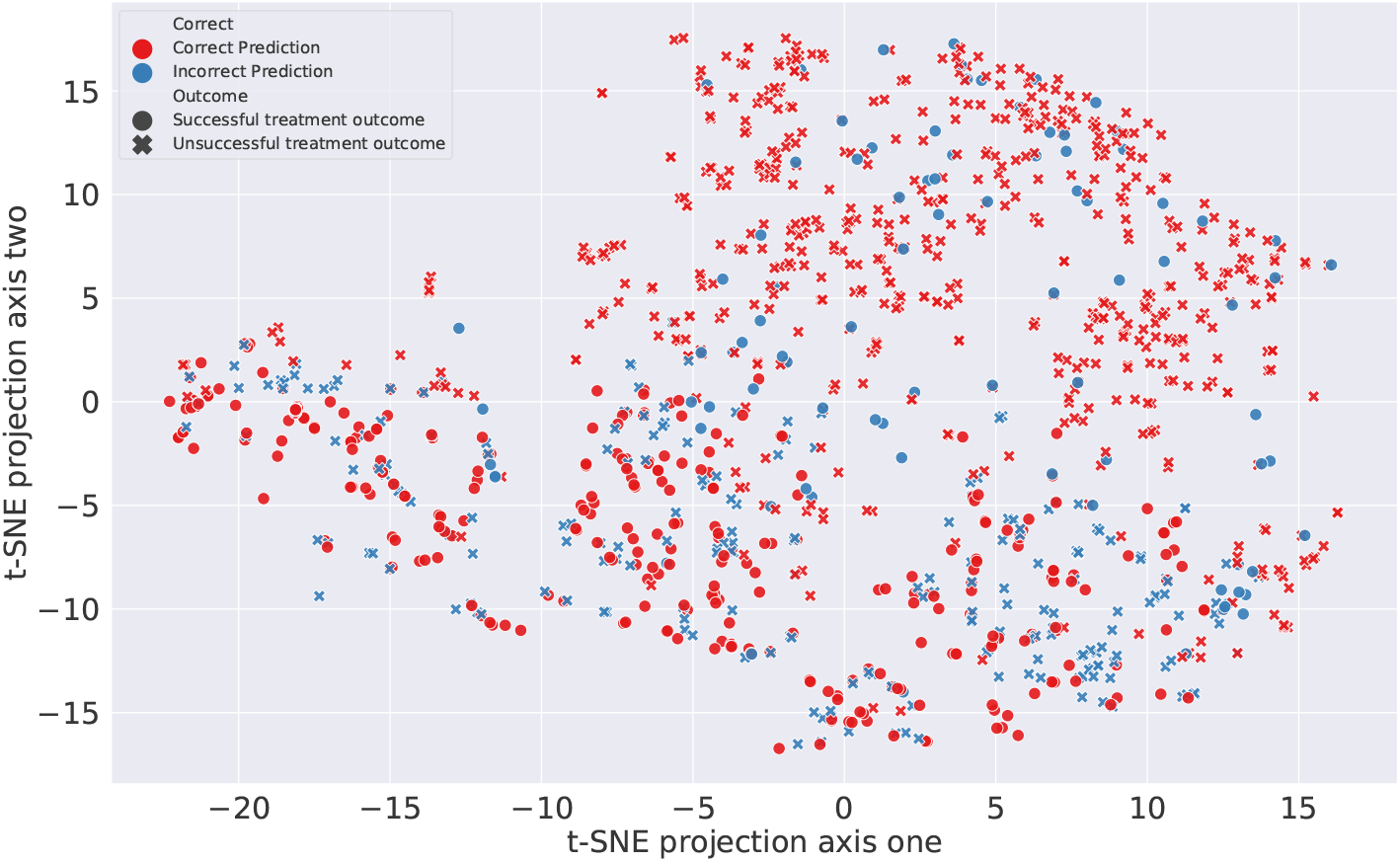
Projection of the different predictions for the treatment outcome of each ASM schedule. There are clusters of incorrect predictions which indicates the complexity of separating the treatment outcomes

## 5. LIMITATIONS AND DISCUSSION

We present a novel approach in adapting a transformer architecture to model epilepsy treatment responses. The approach in this work can greatly advance the application of machine learning in predicting treatment outcome in epilepsy, by using novel deep learning techniques in this area. While previous attempts at the same were done largely through surrogate markers of treatment response,^14, 15, 22^ we have the advantage of being able to train our predictive model using relevant clinical information with known standard treatment outcome measures in two large cohorts of newly diagnosed and treated epilepsy patients.^3, 23^

Although the preliminary results show promise of the transformer architecture being able to predict the outcome of epilepsy treatment, our study has notable limitations such as limited patient information and data. We plan to improve the prediction models by incorporating more clinically relevant input variables for model training such as the latest etiological classification of epilepsy proposed by International League Against Epilepsy (ILAE),^24^ electroencephalographic abnormalities, relevant imaging abnormalities, comorbidities considered during drug selection and concomitant medication use.

In clinical practice, safety of ASM is also factored in while prescribing. An efficacious and tolerable drug may not be the safest to use in a given situation.^21^ For instance, although valproate is efficacious for idiopathic generalised epilepsy, it is not recommended for women of child bearing age due to its teratogenic potential. These sorts of decisions require more nuanced knowledge of balancing pros and cons of selecting a particular regimen. As the model does not naturally account for this limitation, a generalized approach needs to be further investigated to implement these additional considerations.

## 6. CONCLUSION AND CLINICAL OUTCOMES

The usage of an attention-based model to capture relationships within treatment outcomes for epilepsy patients has yielded promising results. The model was able to generalise well to an external cohort without further fine-tuning of the model. In the future, the model can be improved upon with additional variables, which will allow the model to aid clinicians in selecting the most suitable ASM based on its predicted efficacy in an individual.

## Data Availability

Data can be provided to qualified investigators with institutional regulations.

## APPENDIX A. HYPER-PARAMETERS

Figure 4 the list of hyper-parameters that were tuned with with the training/validation split of the Glasgow dataset.

## APPENDIX B. LIST OF VARIABLES

Figure 5 outlines the patient variables that were used to train the model. The ASMs that were used in the Glasgow and Perth datasets are not shown in this section. Further information on the variables can be found in.^21^

## Footnotes

* Detailed explanation of the Glasgow data can be found in^21^

